# Development and validation of a laboratory-developed test on Panther Fusion^®^ system for detection of macrolide resistance in *Bordetella pertussis*

**DOI:** 10.64898/2026.09.22.26363724

**Authors:** Lamali Sadeesh Kumar, Michelle Sait, Benjamin P. Howden, Norelle L. Sherry

**Affiliations:** Department of Microbiology and Immunology, The University of Melbourne at the Peter Doherty Institute for Infection and Immunity, Melbourne, Victoria, Australia; Microbiological Diagnostic Unit Public Health Laboratory, Department of Microbiology and Immunology, The University of Melbourne at the Peter Doherty Institute for Infection and Immunity, Melbourne, Victoria, Australia; WHO Collaborating Centre for Antimicrobial Resistance, The Peter Doherty Institute for Infection and Immunity, Melbourne, Victoria, Australia; Centre for Pathogen Genomics, The University of Melbourne, Melbourne, Victoria, Australia; Department of Infectious Diseases & Immunology, Austin Health, Heidelberg, Victoria, Australia

## Abstract

Macrolides remain the first-line antimicrobial treatment for pertussis, primarily caused by *Bordetella pertussis*. Emergence of macrolide-resistant *B. pertussis* in Australia and globally presents a growing concern for clinical management. As molecular diagnosis predominates for pertussis, limited numbers of cultures are available for antimicrobial susceptibility testing. The A2037G mutation in the 23S rRNA gene is most commonly associated with macrolide resistance in *B. pertussis*, yet no commercial molecular assay is available for its detection. In this study, we adapted and validated a previously published real-time PCR assay as a laboratory developed test (LDT) on the Panther Fusion^®^ system using Open Access−functionality to detect the macrolide resistance-associated mutation in *B. pertussis* from culture isolates and clinical specimens. Total turnaround time was ∼2.5 h for the first result and 2-3 min for subsequent results in the same batch, with an estimated cost of AUD 18 per sample, excluding labour. A total of 30 samples were included in the validation panel to assess assay performance. All isolates and clinical specimens in the validation panel produced results concordant with the expected mutation status, demonstrating 100% accuracy, sensitivity and specificity. The assay demonstrated a limit of detection of 1.5 × 10^3^ CFU/mL (22 CFU/reaction). This Panther Fusion^®^ LDT provides a rapid and fully automated approach for molecular detection of the targeted mutation associated with macrolide resistance in *B. pertussis*. Its integration into routine molecular workflows as a reflex test following identification of *B. pertussis* may facilitate timely identification of resistant infections.

## Introduction

Pertussis (whooping cough) is primarily caused by *Bordetella pertussis* and macrolides such as erythromycin, azithromycin and clarithromycin remain the first-line antimicrobial treatments [1].

Pertussis continues to cause cyclic epidemics in Australia and globally despite the high vaccination rates, with a substantial resurgence reported in 2024 [2, 3]. Macrolide resistance in *B. pertussis* was first reported in 1994 in the United States [4] and remained rare in Australia until its first documented case in 2024 [5]. Macrolide-resistant *B. pertussis* (MRBP) has increasingly been reported in multiple countries following the COVID-19 pandemic, with particularly high resistance rates reported in China [6-8]. Therefore, timely detection and monitoring of MRBP strains have become increasingly important to inform antimicrobial treatment and support resistance surveillance.

Studies have reported an association between macrolide resistance in *B. pertussis* with an A→G substitution at position 2037 of the 23S rRNA gene relative to the *B. pertussis* Tohama I genome (accession no. NC_002929.2), also reported as position 2047 in some literature based on a previous version of the reference genome [9, 10]. *B. pertussis* carries three copies of the 23S rRNA gene, and MRBP strains have been often reported to carry the resistance-associated mutation across all three copies, altering the macrolide-binding site within the ribosomal subunit [3, 9]. Although heterozygous variants at the mutation site have been described, these appear to be uncommon [9, 11]. Phenotypic antimicrobial susceptibility testing (AST) has identified MRBP strains carrying this mutation exhibiting high-level macrolide resistance (minimum inhibitory concentration (MIC) >256 mg/L), with some reports describing microbiological treatment failure [3, 5]. However, the predominance of molecular diagnosis has reduced opportunities to recover *B. pertussis* isolates for phenotypic antimicrobial susceptibility testing. Despite the need for rapid molecular detection of resistance, no commercially available assay currently enables direct detection of macrolide resistance in *B. pertussis*.

In this study, we adapted and optimised a previously published real-time PCR assay on the Panther Fusion^®^ system to enable rapid, automated detection of the A2037G mutation in the 23S rRNA gene associated with macrolide resistance in *B. pertussis* from both culture isolates and directly from clinical specimens. The assay was validated using culture isolates and clinical specimens to assess its performance with the intended use as a reflex test following molecular identification of *B. pertussis*.

## Materials and Methods

### Validation dataset

Thirty samples, comprising reference isolates and clinical specimens, were used to validate the LDT on the Panther Fusion^®^ for detecting macrolide resistance in *B. pertussis*. For this study, *B. pertussis* isolates and clinical specimens containing *B. pertussis* with the A2037G mutation in the 23S rRNA gene were referred to as macrolide-resistant *B. pertussis* (MRBP), while those without the mutation were referred to as macrolide-susceptible *B. pertussis* (MSBP). The validation panel included two reference isolates: one MRBP strain and one MSBP strain (ATCC 9797) and eight nasopharyngeal swab (NPS) specimens collected between 2024 and 2025 comprising four specimens with MRBP and four specimens with MSBP. The clinical specimens were initially reported as PCR-positive for *Bordetella* spp. by primary diagnostic laboratories and referred to MDU PHL for speciation and further genotyping. Speciation performed at MDU PHL confirmed *B. pertussis* in all eight clinical specimens by a real-time

PCR as previously described [12]. Corresponding *B. pertussis* isolates were available from all four clinical specimens with MSBP, however, corresponding isolates were not available from the four clinical specimens with MRBP. The presence or absence of the targeted mutation associated with macrolide resistance was confirmed by whole genome sequencing (WGS) of reference and corresponding culture isolates and by Sanger sequencing of the 23S rRNA gene region encompassing the A2037G mutation in clinical specimens with MRBP. Two NATtrol−Respiratory Panel 2.1 Controls (catalogue number: NATRPC2.1-BIO https://www.zeptometrix.com/au/en/nattroltm-respiratory-panel-21-rp21-controls-12-x-03ml-3084) were also included as commercial controls with Control 2 containing *B. pertussis* A639, a macrolide-susceptible strain, along with other respiratory pathogens. Additionally, 18 NPS specimens tested negative for *B. pertussis* by the BioFire^®^ Respiratory 2.1 (RP2.1) Panel (bioMérieux) (https://www.biomerieux.com/corp/en/our-offer/clinical-products/biofire-respiratory-2-1-panels.html) were included in the validation panel (Table S1).

### Assay design and adaptation to the Panther Fusion^®^ system

A real-time PCR assay previously described for detection of macrolide resistance in *B. pertussis* [2] was adapted for implementation on the Panther Fusion^®^ system (Hologic^®^) using its Open Access−functionality [13]. The assay was initially designed as a duplex real-time PCR targeting the 23S rRNA gene, using a single primer pair and two probes targeting separate regions of the amplified sequence: a 23S rRNA control region and the A2037G mutation site associated with macrolide resistance in *B. pertussis* [2]. However, the 23S rRNA control target was excluded from the final configuration of this assay due to concerns regarding its specificity for *B. pertussis*. The annealing temperature of the initial assay was 48°C, which was outside the performance specifications of the Panther Fusion^®^ instrument. Therefore, the probe targeting the mutation was modified, including incorporation of a locked nucleic acid (LNA) base to increase the annealing temperature while retaining assay specificity. Primer and probe sequences used in this study are listed in Table 1.

**Table 1:**
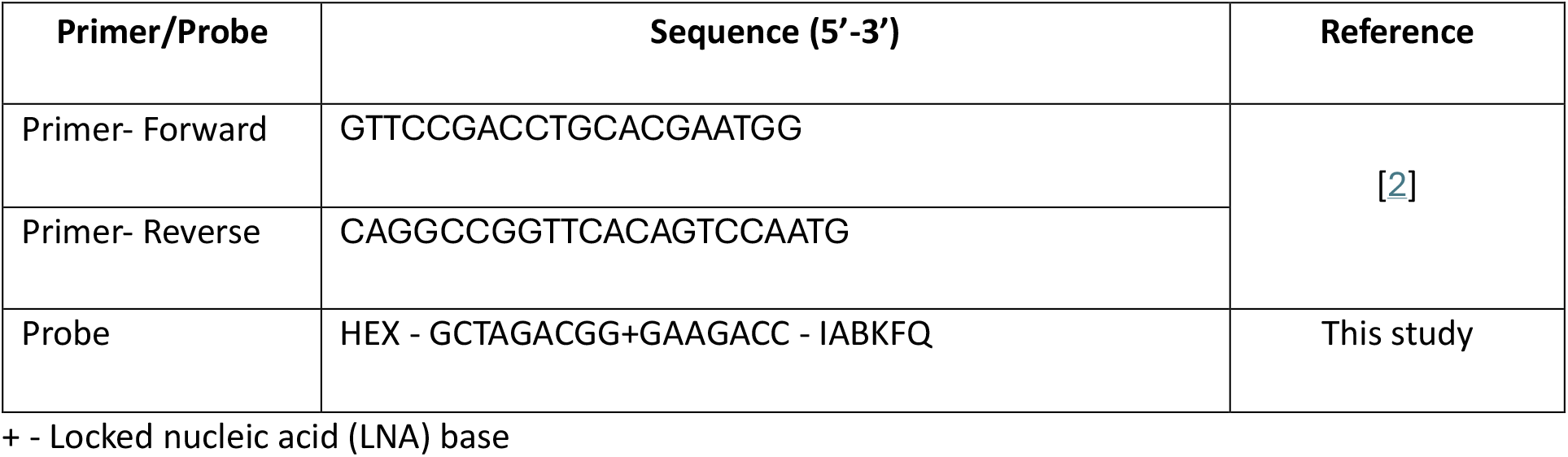
The primer and probe sequences used in this study.

### Panther Fusion^®^ Laboratory Developed Test

Reference culture strains or 500 µL of clinical specimens were inoculated into Panther Fusion^®^ Specimen Lysis Tubes (Hologic^®^ PRD-04339) and 360 µl sample extracted onboard the Hologic Panther Fusion^®^ system using the FCR-S/FER-S extraction kit (Hologic^®^ PRD-04331) and eluted into a volume of 50 µL.

Primer and probe reagent (PPR) mixture consisted of 2 mM MgCl_2_, 60 mM KCl, 10 mM Tris buffer, 0.5 µM each primer and probe and 0.2 µM each Panther Fusion^®^ IC primer and probe. PCR was performed on the Panther Fusion^®^ system using 5 µL of extracted DNA and Open Access™ RNA/DNA Enzyme Cartridge (Hologic^®^ PRD-04303). PCR amplification consisted of an initial holding stage at 95°C for 2 min, followed by 45 cycles at 95°C for 8 s and 60°C for 30 s, with fluorescence measured during the 60°C step. Projected runtime for the assay was 54 minutes and 38 seconds. Data analysis was performed onboard using Curve Correction Method Option 2 and enhanced resolution disabled. Baseline correction was enabled for all analytes, with analysis commencing at cycle 10 and a slope limit of 50. The targeted mutation and IC were detected using HEX and Quasar 705 channels, respectively.

Results were interpreted onboard the Panther Fusion^®^ system using optimised criteria configured in myAccess−software (v3.0.1.19). For the targeted mutation, the fluorescence threshold was set at 400 RFU with a minimum slope of 100 and for IC, the fluorescence threshold was set at 500. Sample validity was determined using IC, with samples considered valid when targeted mutation and/or IC was detected.

### Evaluation of Performance Characteristics

Results obtained from the validation panel were used to compare against expect results and determine assay accuracy, sensitivity, specificity and predictive values (positive and negative). For Limit of Detection (LoD) testing, three replicates of a 0.5 McFarland suspension of the MRBP reference isolate, corresponding to ∼ 1.5 × 10^8^ CFU/mL were prepared and serially diluted to concentrations as low as 1.5 CFU/mL for testing. Data from the dilution series were also used to determine assay precision (repeatability and reproducibility), measuring interval and PCR efficiency.

## Results

The MRBP reference isolate and all four NPS specimens confirmed to contain MRBP tested positive for the A2037G mutation in the 23S rRNA gene with the Panther Fusion^®^ LDT. The MSBP reference isolate and all four NPS specimens confirmed to contain MSBP tested negative for the mutation. Both NATtrol−Respiratory Panel 2.1 (RP2.1) Controls (Control 1 and 2) and all NPS specimens confirmed negative for *B. pertussis* (*n*=18) were negative for the targeted mutation. Valid internal control results were obtained for all samples with no evidence of assay inhibition. Overall, all samples in the validation panel were concordant with the expected results for detecting A2037G mutation in the 23S rRNA gene, with four true-positive and 26 true-negative results and no false-positive or false-negative results. No cross-reactivity with other respiratory pathogens or commensal flora was observed in clinical specimens negative for *B. pertussis*. Accordingly, sensitivity, specificity, positive predictive value and negative predictive value were all calculated as 100%.

For detection of A2037G mutation in the 23S rRNA gene in MRBP, a maximum Ct of 37 was established during assay optimisation to mitigate false-positive detection at later amplification cycles. Following the established Ct threshold, LoD testing demonstrated a detection limit of 1.5 × 10^3^ CFU/mL (22 CFU/reaction) for the targeted mutation. Precision (repeatability and reproducibility) at the LoD was 100% with all three replicates detected and a mean Ct of 36.6. The assay consistently detected the targeted mutation across the tested concentration range of 1.5 × 10^3^ to 1.5 × 10^7^ CFU/mL (22 to 2.2 × 10^5^ CFU/reaction), with all three replicates detected at each concentration (Table 2). PCR efficiency was calculated from mean Ct values across the detected concentration range and was 100.8%, within the acceptable range of 90-110%.

**Table 2.** Limit of detection (LoD) testing of the Panther Fusion^®^ LDT.

| MRBP concentration in original specimen (CFU/mL) | CFU/reaction | Mean Ct – A2037G in the 23S rRNA gene | Number of replicates detected |
| --- | --- | --- | --- |
| $1.5 \times 10^7$ | $2.2 \times 10^5$ | 23.1 | 3/3 |
| $1.5 \times 10^6$ | $2.2 \times 10^4$ | 26.8 | 3/3 |
| $1.5 \times 10^5$ | $2.2 \times 10^3$ | 30.0 | 3/3 |
| $1.5 \times 10^4$ | 223 | 32.9 | 3/3 |
| $1.5 \times 10^3$ | 22 | 36.6 | 3/3 |
| 150 | 2 | neg | 0/3 |
| 15 | 0 | neg | 0/3 |

The complete workflow, including automated nucleic acid extraction, provided the first result in ∼2.5 hours, with subsequent results generated every 2-3 minutes within the batch. The estimated cost was AUD 18 per sample, excluding labour costs.

## Discussion

Macrolide-resistant *Bordetella pertussis* (MRBP) has been increasingly reported in Australia and internationally during recent years [2, 3, 5-7]. Given the scarcity of culture isolates, implementation of culture-independent methods to detect macrolide resistance is important for routine diagnostics as macrolides remain the first-line treatment for pertussis. In this study, we adapted a previously published real-time PCR assay and optimised it as a laboratory-developed test on the Panther Fusion^®^ system to detect the A2037G mutation in 23S rRNA gene associated with macrolide resistance in *B. pertussis* directly from clinical specimens as well as from culture isolates.

The association between the A2037G mutation in the 23S rRNA gene and macrolide resistance in *B. pertussis* has been previously described [9], leading to the development of molecular assays for detection of this resistance-associated point mutation. Such assays include conventional gel-based PCR and real-time PCR [11, 14], which typically require separate nucleic acid extraction and PCR amplification steps. In this study, the assay was adapted and optimised for the Panther Fusion^®^ system integrating automated nucleic acid extraction, PCR amplification and result interpretation within a single platform. Utility of the Panther Fusion^®^ system provides a practical advantage for diagnostic laboratories already using automated molecular workflows by reducing manual processing and facilitating incorporation of macrolide resistance testing of *B. pertussis* into routine diagnostic workflows. The assay achieved a turnaround time of 2.5 hours for the first result with subsequent results available within 2-3 min in the same batch, while substantially reducing hands-on processing compared with conventional PCR workflows. However, this assay may also be adaptable to off-board real-time PCR platforms, providing additional flexibility for laboratories without a Panther Fusion^®^ system.

An important consideration is the specificity of resistance testing based on the 23S rRNA target. The A2037G mutation is in domain V of the 23S rRNA gene, which shares sequence similarity among *Bordetella* species, and *B. pertussis* contains three copies of the 23S rRNA gene [9]. Therefore, the Panther Fusion^®^ LDT should be implemented as a reflex test following the detection of *B. pertussis* in clinical specimens by a commercial or an in-house PCR assay. Moreover, careful interpretation of amplification signals is important for reliable detection of the targeted mutation. In this assay, the established Ct <37 threshold provides a defined cut-off to support specificity and reduce the potential for false-positive results.

The validation panel for this assay comprised a small set of MRBP samples, including one MRBP isolate (Reference) and four clinical specimens containing MRBP. Given the low prevalence of macrolide resistance in Australia, approximately 4% in 2024 [2], obtaining additional resistant isolates and MRBP-positive clinical specimens for validation was challenging. While all macrolide-resistant and macrolide-susceptible *B. pertussis* isolates and clinical specimens produced the expected results, with 100% sensitivity and specificity, evaluation using an extended dataset with more MRBP-positive isolates and clinical specimens would provide further confidence in assay performance.

Detection of the A2037G mutation in the 23S rRNA gene provides genotypic evidence of macrolide resistance but does not directly establish phenotypic resistance. Nevertheless, some studies have reported *B. pertussis* strains exhibiting phenotypic macrolide resistance in the absence of the A2037G mutation in the 23S rRNA gene [14, 15]. Hence, culture and phenotypic antimicrobial susceptibility testing should be performed where feasible to confirm resistance, support surveillance of macrolide-resistant strains and identify emerging resistance mechanisms in *B. pertussis*.

In conclusion, the Panther Fusion^®^ LDT provides a rapid and automated approach for detecting the A2037G mutation in the 23S rRNA gene associated with macrolide resistance in *B. pertussis* directly from clinical specimens and culture isolates. Its integration into routine molecular diagnostic workflows may facilitate timely identification of macrolide-resistant pertussis infections, inform treatment decisions and support ongoing antimicrobial resistance surveillance.

## Data Availability

All data produced in the present study are available upon reasonable request to the authors

## Author statements

### Author contributions

L.S.K. – conceptualization, wet-lab work, data curation and analysis, writing original draft. M.S. – conceptualization, wet-lab work, data analysis, writing-review and editing. N.L.S. – conceptualization, writing-review and editing, funding. B.P.H. – conceptualization, writing-review and editing, funding.

### Ethical approval

Ethical approval for collection of clinical specimens was obtained from the University of Melbourne Human Research Ethics Committee under the project “Pathogen Recovery in Human Clinical Samples” (Reference Number: 2021-22158-20892-3).

## Acknowledgements

We acknowledge and thank the diagnostic laboratories in the state of Victoria for referring clinical specimens that were PCR-positive for *Bordetella* spp. to Microbiological Diagnostic Unit Public Health Laboratory (MDU PHL) for this study. The authors gratefully acknowledge Rebecca J. Rockett (PhD) and staff from the Western Sydney Local Health District, NSW, Australia for providing the macrolide-resistant *B. pertussis* reference isolate.

